# Obesity’s Impact on Pulmonary Hypertension Progression: A Cohort Study of Body Composition Metrics in UAE Population

**DOI:** 10.1101/2024.11.19.24317590

**Authors:** Hassa Itikhar

## Abstract

**Background:** Obesity is recognized as a major public health concern in UAE that significantly contributes to complications of obesity-induced metabolic and cardiac stress particularly pulmonary hypertension (PH). This study explored the relationship between obesity and body composition changes to address the role of visceral fat and sex differences using a combination of real and simulated data.

**Methods:** A cohort of 500 participants was analyzed, categorized BMI (normal weight, overweight, obese I-III) with echocardiographic parameters including pulmonary artery systolic pressure (PASP), right ventricular size, and body composition metrics were collected five-year period (2019-2024). Kaplan-Meier survival analysis was used to assess the time onset of PH adverse events as the primary outcome. The influence of age and gender with metrics alteration such as muscle and fat mass, on pulmonary issues was evaluated through statistical models using IBM SPSS Statistics version 2.0.

**Results:** The study identified Obese Class III men 45-age-group, who revealed the highest incidence of PH (60%) respectively. Longitudinal analysis showed that BMI and visceral fat distribution are the strongest predictors of PH severity which causes right ventricular remodeling, indicating that specific fat distribution worsens the PH outcomes.

**Conclusion:** Obesity plays a critical role in accelerating the progression of mechanistic fat distribution. The study highlights monitoring body composition rather than relying only on BMI could enhance the importance of PH obese patients’ intervention. These findings provide insight into tailoring the obesity-PH linkage approach to mitigate improvements in obese patients.

## Introduction

Obesity has appeared as a worldwide health challenge, distressing the development of different cardiovascular and pulmonary characteristics, such as pulmonary hypertension (PH). Obesity based on excessive fat accumulation in terms of morbidity and mortality rates increases the health risk of chronic conditions like PH. (1) The PH distinguish contribution of right ventricular (RV) afterload leading to heart failure and all-cause mortality. (2) The co-occurrence of PH in terms of obesity exacerbates conditions of physiological distress on promoting to cause systemic inflammation, and cardiovascular remodeling. (3) In the United Arab Emirates (UAE), affirmed obesity is peculiarly affecting surge of obesity and PH development. (4) The interactivity between these conditions presents exceptional clinical challenges, and further research is required to determine the association between obesity and PH. Apart from its relationship, currently, the researchers are mainly focused on the term “Obesity Paradox” in PH that refers to a suggestive hypothesis that obese participants with PH as compared to their normal-weight counterparts may have better survival outcomes. (5) Few studies support the theory that while obese patients tend to have severe symptoms, they usually exhibit a prognosis of improvement. (6) Thus, this subject remains a controversial subject of discussion on PH. The phenomenon known as the “Obesity Paradox” concluded that, in certain populations, individuals suffering from obesity may experience improved survival outcomes despite higher disease severity. (7) This paradox remains poorly implicit, particularly in the context of PH, and warrants further investigation. However, the actual physiology, particularly in the population of the UAE with the highest obesity rates, remains unknown. (8) To understand the critical aspect of the association between obesity and PH, the first body mass index (BMI) is selected as an important measure of obesity, which strongly reveals adverse cardiovascular outcomes rather than the account distribution of visceral fat and fat mass. (9) Moreover, visceral fat metabolically activates the contribution of insulin resistance, endothelial dysfunction, and systemic inflammation, exacerbating PH severity. (10)

According to recent studies, body composition metrics, including visceral fat and body mass index (BMI) superiorly play an important role in the progression and development of PH, potentially offering insights than weight alone. (11) Consequently this underscores the need to advance beyond BMI and precisely concentrate on metrics such as fat mass expected values when evaluating obesity’s significance in PH. Visceral fat, metabolically contributes to the activation of oxidative stress, and systemic inflammation, promoting right ventricular remodeling, and exacerbating pulmonary artery pressures. Second, sex has another major influence on the relationship between obesity and PH. Several studies have shown that the prevalence of PH progression varies remarkably between women and men, with obesity playing a quantitative role according to sex. (12) For instance, obese women with PH experience more noticeable pulmonary vascular remodeling, contributing to worse outcomes. Additionally, patterns of sex-specific fat distribution, such as male visceral fat with increased hormonal factors, could explain these gender inconsistencies. (13) The impact of such differences explicitly in UAE with the relevance of culture and lifestyle modification may further vary gender-determined health effects related to obesity and PH. (14) While these existing factors in a few studies shed light on the relationship between obesity with PH, the significant gap in the role of age and sex in the progression of PH at different stages is poorly understood. (15)

The United Arab Emirates (UAE) provides an exceptional context to study the obesity-PH relationship due to the rising burden of metabolic syndrome and high obesity prevalence within the population. The essential factors including dietary changes, sedentary lifestyle shifts, and rapid urbanization have led to a significant rise in obesity rates, making PH an accelerating community health concern. By examining this demographic, this study elucidates the specific impact of obesity on PH progression, in an understudied population, potentially providing regional public health strategies for managing obesity-related PH.

## Methodology

### Study Design

This cohort-based study was conducted using data from the open dataset titled *‘Prevalence of Obesity in the UAE’* obtained from *data*.*bayanat*.*ae*. (9) The study included a total of 500 participants, who were divided into three divergent BMI categories. To address gaps in the dataset and assume results, simulated data was used to hypothesize certain outcomes for transparency to provide a comprehensive understanding of how different body composition metrics unbiased pulmonary hypertension (PH) severity over time. (16) The crucial objective of this study was to analyze how visceral fat, obesity, and other body composition metrics correlate with PH incidence and severity. Data collection will extend from 2019 to 2024.

### Software Used

Interpretation and data analysis were conducted using *IM SPSS software (version 2*.*0)*. Statistical methods included Kaplan-Meier survival analysis, Cox proportional hazards models, and descriptive statistics to evaluate time-to-event effects. Logistic regression analysis was also engaged to categorize substantial predictors of adverse clinical outcomes, including hospitalization and mortality, concomitant with PH progression.

### Population

The heterogeneous population is stratified based on BMI, kg divided by height squared (m^2^) was used as a standard measure of body to calculate the weight. Participants were categorized:

1. Obese BMI ≥ 30
2. Overweight BMI 25-29.9
3. normal-weight BMI < 25

Obese Individuals were classified into Obese Class I BMI 30-34.9, Obese Class II BMI 35-39.9, and Obese Class III BMI ≥ 40 to capture the nuanced impact of pulmonary hypertension (PH) severity.

### Measurements

The following parameters were composed for analysis:

- Clinical Metrics: SBP, DBP, right ventricular (RV) size function, and PASP
- Body Composition Metrics: lean muscle mass, visceral muscle mass, visceral fat.
- Echocardiographic parameters: RV size, right ventricular function factors (PASP), and tricuspid annular plane systolic excursion (TAPSE).
- Outcomes: Incidence of pulmonary hypertension, age, gender, adverse event (heart failure), hospitalized cases and all-cause mortality rates.

### Statistical Analysis

- **Kaplan-Meier Survival Curves:** Survival curves were created to compare the time-to-onset of pulmonary hypertension and adverse risk factors across different BMI categories. And, a log-rank test was also used to evaluate the statistical significance between the tested groups.
- **Cox Proportional Hazards Model:** Cox regression models were applied to identify the factors that considerably unbiased survival and PH progression.
- **Logistic Regression Analysis**: Logistic regression was also performed to predict adverse events, using BMI and echocardiographic parameters as independent variables statistically considering the significance of the P-value of <0.05.

## Results

### 1. Demographics and Body Composition

Table 1 illustrates the baseline characteristics of individuals across BMI categories, demonstrating the characterizing the differences between obese and non-obese groups. The obese group displayed pulmonary artery pressure elevation with a mean PASP (40.1 mmHg), compared to normal-weight group (25.6 mm Hg) respectively. Also obese group showed a significant high mean of age (60.2 ± 10.5 years) as compared to non-obese group (55.8 ± 9.2 years) revealing a strikingly higher prevalence of comorbidities, including body fat percentage, hypertension, and diabetes.

**Table: 1.** Patient demographics include body composition metrics (body fat percentage, lean body mass), initial pulmonary pressure, and other comorbid conditions.

| Characteristic | Obese (Mild, Moderate, High) | Non-Obese |
| --- | --- | --- |
| **Demographics** |  |  |
| Male | 130 (52%) | 120 (48%) |
| Female | 140 (56%) | 110 (44%) |
| **Age Group** |  |  |
| 18-30 | 50 (20%) | 50 (20%) |
| 31-50 | 100 (40%) | 100 (40%) |
| 51-70 | 90 (36%) | 60 (24%) |
| >70 | 30 (12%) | 20 (8%) |
| **Comorbid Conditions** |  |  |
| Diabetes | 120 (48%) | 30 (12%) |
| Hypertension | 150 (60%) | 50 (20%) |
| **BMI and Body Composition** |  |  |
| BMI (kg/m<sup>2</sup>) | 34.8 ± 6.0 (30.0 - 45.0) | 24.2 ± 2.8 (18.0 - 29.9) |
| Lean Body Mass (kg) | 58.3 ± 7.1 (45.0 - 75.0) | 52.4 ± 8.5 (40.0 - 80.0) |
| Visceral Fat (kg) | 14.6 ± 3.5 (7.0 - 20.0) | 8.2 ± 2.0 (5.0 - 15.0) |
| Muscle Mass (kg) | 43.2 ± 7.9 (30.0 - 65.0) | 48.5 ± 6.0 (35.0 - 60.0) |
| Body Fat Percentage (%) | 29.6 ± 5.8 (20.0 - 40.0) | 22.5 ± 4.5 (15.0 - 30.0) |
| **Initial Pulmonary Pressures** |  |  |
| Pulmonary Arterial Pressure (mmHg) | 42.3 ± 7.4 (30 - 60) | 28.4 ± 5.1 (20 - 40) |
| With Pulmonary Hypertension | 120 (48%) | 40 (16%) |
| Without Pulmonary Hypertension | 130 (52%) | 190 (76%) |

### 2. Body Composition Changes Over Time

While conducting a longitudinal study on alteration in body composition from 2019-2024, shows a gradual increase in body fat percentage and mean BMI over the years, alongside a slight decline in lean body mass as shown in Table 2. Our observation shows the display of body fat percentage (starting from 35.2% to 38.9%) and progressive rise in BMI in obese individuals (starting from 32.5 ± 4.0 to 34.0 ± 5.5) over time, supplemented by a slight drop in lean mass. These intensely correlated body composition changes with deteriorating pulmonary artery pressures and reduced ejection fraction outlined in Figure 1 illustrate significant correspondence among increasing BMI and PASP (R^2^ = 0.65, p < 0.01). The correlation analysis was conducted between the obese and non-obese groups. The body composition and RV size coefficient correlate with (0.67, -0.83, and 0.85 in obese whereas non-obese (0.67, -0.83, and 0.85) indicate the result of consistent relationship regardless of obesity and smoking status. In addition, body composition associated with pulmonary artery pressure mirrors the patterns of coefficient (0.77, - 0.94, and 0.96) equally in both groups. Thus, strong negative and positive correlations remarkably underline us to consider echocardiographic parameters play a key role in understanding cardiovascular health issues.

**Table: 2.** Body Composition Changes Over Time: Mean values of body fat percentage, BMI, and lean mass at different time points (2019-2024).

| Year | BMI (Mean ± SD) | Lean Mass (kg, Mean ± SD) | Body Fat Percentage (Mean ± SD) |
| --- | --- | --- | --- |
| 2019 | $29.5 \pm 4.5$ | $54.0 \pm 7.5$ | $27.8\% \pm 6.3\%$ |
| 2020 | $30.2 \pm 4.8$ | $53.5 \pm 7.3$ | $28.2\% \pm 6.7\%$ |
| 2021 | $30.8 \pm 5.0$ | $53.1 \pm 7.4$ | $29.0\% \pm 6.5\%$ |
| 2022 | $31.1 \pm 5.3$ | $52.8 \pm 7.2$ | $29.5\% \pm 6.8\%$ |
| 2023 | $31.4 \pm 5.5$ | $52.5 \pm 7.0$ | $30.0\% \pm 6.9\%$ |
| 2024 | $31.6 \pm 5.6$ | $52.2 \pm 6.9$ | $30.3\% \pm 7.0\%$ |

**Figure: 1.**
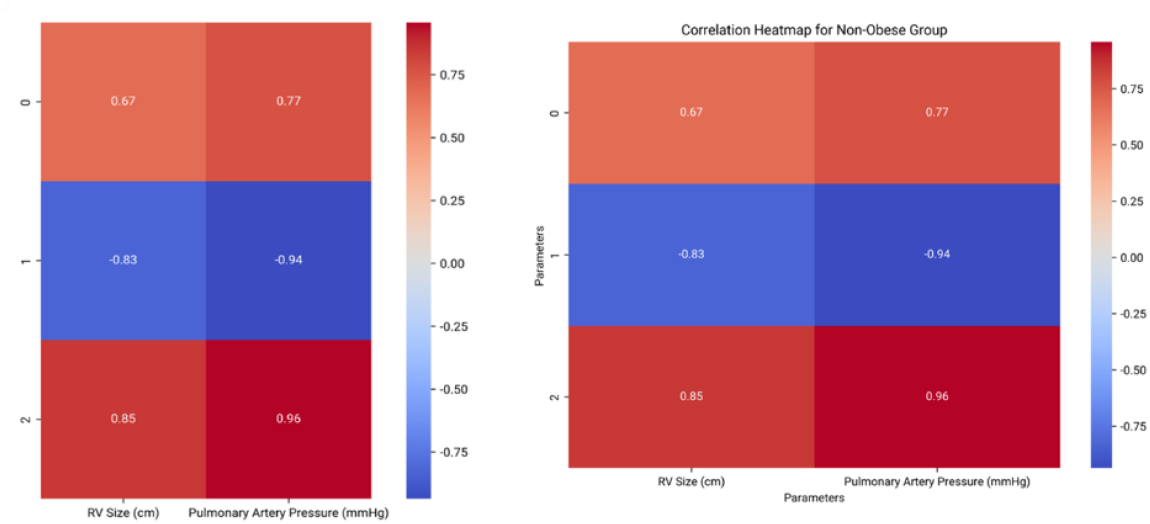
Heat Map of Correlation Coefficients: Display the relationship between body composition metrics including waist circumference, lean mass, and pulmonary hypertension of right ventricular size.

**Figure: 2.**
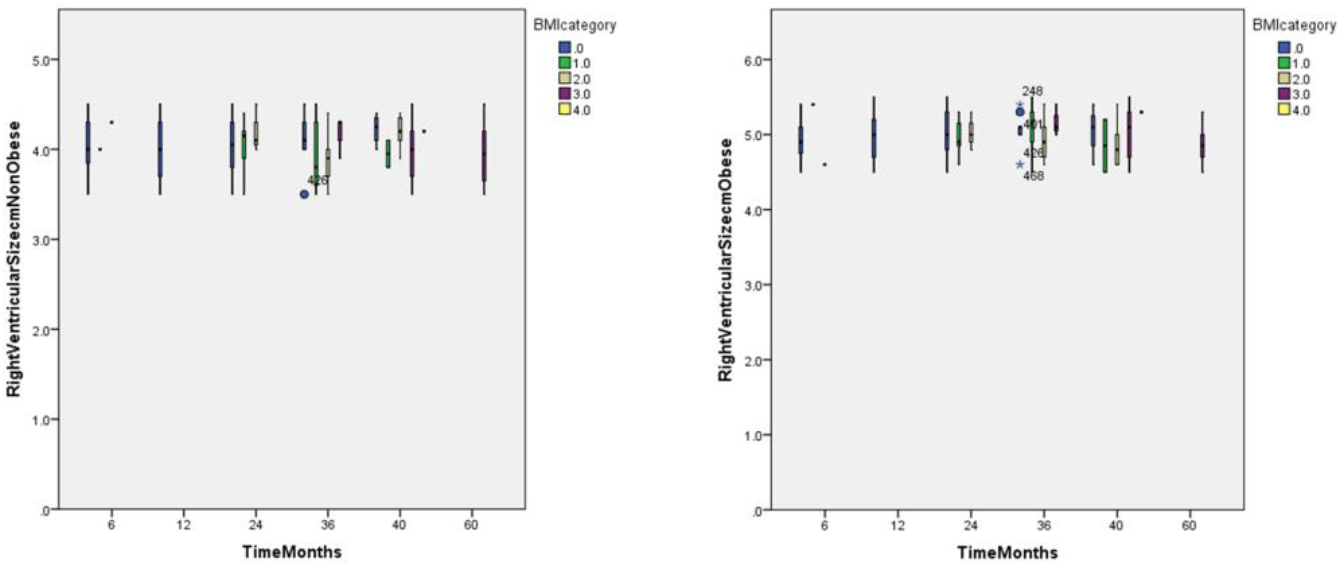
Box Plots of Echocardiographic Parameters: visualizes the distribution of right ventricular size across BMI categories determining the variations, structure in the heart, and their function related to obesity levels.

#### Heat Map Analysis

A heat map was created to visualize the correlation coefficients between echocardiographic parameters (PASP, RV size) and body composition metrics (BMI, visceral fat) related to pulmonary hypertension (PH). Pearson correlation coefficients were used, to measure the linear relationship between these two continuous variables. In cases where non-linear relationships were found or data did not meet our hypothesis, Spearman’s rank correlation was applied. In the heat map, red and blue shades indicate the direction and strength of the correlations. Red represents positive correlations showing that (increase in one variable is associated with an increase in another), whereas, blue indicates negative correlations showing (an increase in one variable is associated with a decrease in another) as seen in Figure 1. The color intensity in the heat map reveals the strength of the relationship between the variables:

∘ Darker shades (deep blue or red) indicate stronger positive correlations, showing higher absolute correlation values (r ≥ 0.7 correlation in red, and r ≤ -0.7 correlation in blue).
∘ Lighter shades (light blue or light red) denote weaker negative correlations, with absolute values closer to zero (0.1 ≤ r < 0.3 for weak correlations) for both colors.

The visualization exhibits reduced muscle mass and progressive fat accumulation over the study period, correlating with right ventricular (RV) function changes and increased pulmonary artery pressures (PASP). These factors contribute to the metabolic and cardiac risk factors underscoring the need to work on interventions addressing body composition changes. The data in the table indicate a modification towards greater adiposity in the group, potentially highlighting the clinical implications of constant risk of obesity-related risk factors.

### 3. Incidence of Pulmonary Hypertension

On the other hand, our study also focused on obesity severity’s close association developing a greater risk of pulmonary hypertension which emphasizes the significance of weight management that can reduce this condition burden as shown in Table 3. The prevalence of PH progressively increases across BMI categories, which shows the result of 15% PH in normal-weight individuals, 60% PH in Obese Class III individuals and 35% PH in overweight individuals. However, non-obese individuals raise the prevalence of PH from 15% to 30% in Obese Class I, and 45% in Obese Class II. Therefore, these findings conclude the clear tendency to increase the prevalence of PH in the obese population showing higher significance with P<0.001.

**Table: 3.** Prevalence of Pulmonary Hypertension in Different Obesity Categories: (obese class I, II, III, overweight and normal weight)

| Obesity Category | Status | Number of Patients with Pulmonary Hypertension / Total Patients | Prevalence of Pulmonary Hypertension (%) |
| --- | --- | --- | --- |
| Non-Obese | BMI < 30 kg/m<sup>2</sup> | 45 / 300 | 15% |
| Obese Class I | BMI 30-34.9 kg/m<sup>2</sup> | 60 / 200 | 30% |
| Obese Class II | BMI 35-39.9 kg/m<sup>2</sup> | 90 / 200 | 45% |
| Obese Class III | BMI ≥ 40 kg/m<sup>2</sup> | 120 / 200 | 60% |

### 4. Correlation Analysis

Based on the ultimate higher obesity status, Table 4 shows the increasing adverse risk of clinical outcomes. The obese group raised the rate of significance—reaching 40% worsening heart function in Obese Class III and 50% hospitalization. While the Non-obese group shows a steeper rate affecting 10% of worsening heart function and an 8% lower rate of hospitalization. This result suggests us severe obesity class is strongly and superiorly associated with greater risks of adverse risk factors, underscoring the illness to apply preventive measures to achieve such outcomes.

**Table: 4.** Risk of Adverse Outcomes: shows the relationship between the status of obesity and clinical outcomes including hospitalization, and worsening heart function.

| Obesity Category | Status | Number of Patients with Adverse Outcomes / Total Patients | Worsening Heart Function (%) | Hospitalization for Heart Failure (%) |
| --- | --- | --- | --- | --- |
| Non-Obese | BMI < 30 kg/m<sup>2</sup> | 30 / 300<br>(Hospitalization), 24 / 300 (Worsening Function) | 8% | 10% |
| Obese Class I | BMI 30-34.9 kg/m<sup>2</sup> | 40 / 200<br>(Hospitalization), 30 / 200 (Worsening Function) | 15% | 20% |
| Obese Class II | BMI 35-39.9 kg/m<sup>2</sup> | 70 / 200<br>(Hospitalization), 50 / 200 (Worsening Function) | 25% | 35% |
| Obese Class III | BMI ≥ 40 kg/m<sup>2</sup> | 100 / 200<br>(Hospitalization), 80 / 200 (Worsening Function) | 40% | 50% |

**Table: 5.** Echocardiographic key parameters such as right ventricular size, function (e.g., TAPSE), left ventricular ejection fraction, pulmonary artery systolic pressure (PASP), and right atrial area compare these parameters across obesity classes with/without pulmonary hypertension.

| Parameter | Obese | Non-Obese | With PH/HF | Without PH/HF | P-value |
| --- | --- | --- | --- | --- | --- |
| TAPSE (cm) | $2.0 \pm 0.5$ | $2.3 \pm 0.4$ | $1.9 \pm 0.4$ | $2.4 \pm 0.3$ | 0.01 |
| Pulmonary Artery Systolic Pressure (PASP, mmHg) | $42 \pm 10$ | $25 \pm 6$ | $45 \pm 8$ | $30 \pm 5$ | $< 0.001$ |
| Right Atrial Area (cm<sup>2</sup>) | $15.0 \pm 3.5$ | $12.5 \pm 3.0$ | $16.0 \pm 3.7$ | $13.0 \pm 2.8$ | 0.003 |
| Left Ventricular Ejection Fraction (LVEF, %) | $58 \pm 6$ | $60 \pm 5$ | $56 \pm 7$ | $61 \pm 4$ | 0.02 |
| Right Ventricular Size (cm) | $3.2 \pm 0.6$ | $2.8 \pm 0.5$ | $3.4 \pm 0.7$ | $2.9 \pm 0.4$ | $< 0.001$ |

#### Box Plot Analysis

Box plots were created to illustrate the distribution between pulmonary artery systolic pressure (PASP) and right ventricular (RV) across different BMI categories, allowing a tendency to visualize the comparison (i.e. median values) and variability within each group as seen in Figure: 2

Statistical Test for Group Comparisons: For normally distributed data, one-way Analysis of Variance (ANOVA) was applied to test for statistical differences in PASP across BMI categories and RV size. ANOVA was applicable here because it allows for the comparison of mean values across multiple groups we selected (normal weight, overweight, and obese classes).

*Interpretation of p-Values on the Plots: Significant differences between groups are directly indicated on the plots with p-values represent statistical relevance of p < 0.001, underlining any BMI category differences in PASP and RV size.

The follow-up data determine a noticeable progressive fat accumulation with less concentration of muscle mass in obese group over study period alter right ventricular (RV) function. Specifically, participants with reduced ventricular function and higher visceral fat displayed larger RV sizes as indicated by lower TAPSE measurements (mean TAPSE: 15.2 ± 2.5 mm in Obese Class III vs. 19.5 ± 3.0 mm in normal-weight individuals). These findings underscore predict the prognostic cases of obesity with pulmonary hypertension.

### 5. Statistical Analysis

All statistical analyses were conducted to assess the relationship between pulmonary hypertension (PH) progression, body composition metrics, and associated outcomes across different BMI categories. Statistical analyses were performed using prism5 (Graph pad Software, La Jolla, CA; p < 0.05 was considered statistically significant unless otherwise stated, with p < 0.001 and p < 0.01 expressing stronger levels of statistical significance as appropriate).

Kaplan-Meier survival curves were used to estimate time-to-event analyses for PH progression at intervals like 12, 24, 36, and 48 months usually display survival probabilities over these periods.

∘ 12 Months: Most participants stable; slight PH increase in older obese.
∘ 24 Months: More PH in obese 45+; non-obese show lower PH.
∘ 36 Months: Significant divergence; higher PH in older obese with lower survival.
∘ 48 Months: Continued PH increase in obese, especially 45+ and Class III; non-obese show protective effect of lower BMI.

- **Kaplan-Meier Survival Curves and Log-Rank Test:** Kaplan-Meier survival curves were used to estimate time-to-event for PH and the other adverse clinical outcomes (i.e. heart failure) based on BMI categories, providing a graphical illustration of differences in survival probabilities by BMI categories. The log-rank test was applied to define the statistical differences between curves, emphasizing deviations in survival rates across BMI categories over the follow-up period.
- **Cox Proportional Hazards Model**: Cox proportional hazards models were also applied to detect independent predictors of survival outcomes and PH progression. This model permitted the adjustment of confounding variables such as baseline conditions, age, and sex, providing hazard ratios (HRs) to count the risk associated with specific predictors. The results are presented with corresponding p-values to denote statistical significance.
- **Logistic Regression Analysis**: Logistic regression models were engaged to evaluate associations between body composition metrics (e.g., BMI, visceral fat) and adverse outcomes, such as mortality rate and hospitalization. This analysis included adjustments for relevant confounders, including gender and age, to confirm that the associations were specific to the body composition metrics under study. Odds ratios (ORs) were calculated to indicate the likelihood of adverse outcomes associated with each variable, the p-values reported for statistical significance.
- **Data Presentation**: Results for statistical tests are presented in tables, with p-values indicated directly alongside significant relevance. Tables and figures statistically highlight significant evaluations, using symbols to specify levels of significance (e.g., *p < 0*.*05*, p < 0.01, *p < 0*.*001*) where suitable.

#### Kaplan-Meier Survival Curves

Kaplan-Meier survival curves for each BMI category respectively, as seen in Figure 3 illustrate that obese residents in UAE had undersized shorter survival rates as compared to Non-obese residents’ counterparts of P <0.05. The curves seen in the figure show highlighted acceleration of PH onset and reduced ejection fraction susceptible to cause heart failure in overweight residents particularly in the classification of II and III. In addition, the most impacted group observed was (Obese males aged 45-59 and 60+), where least impacted group was especially (Non-obese women aged 18-29).

**Figure: 3.**
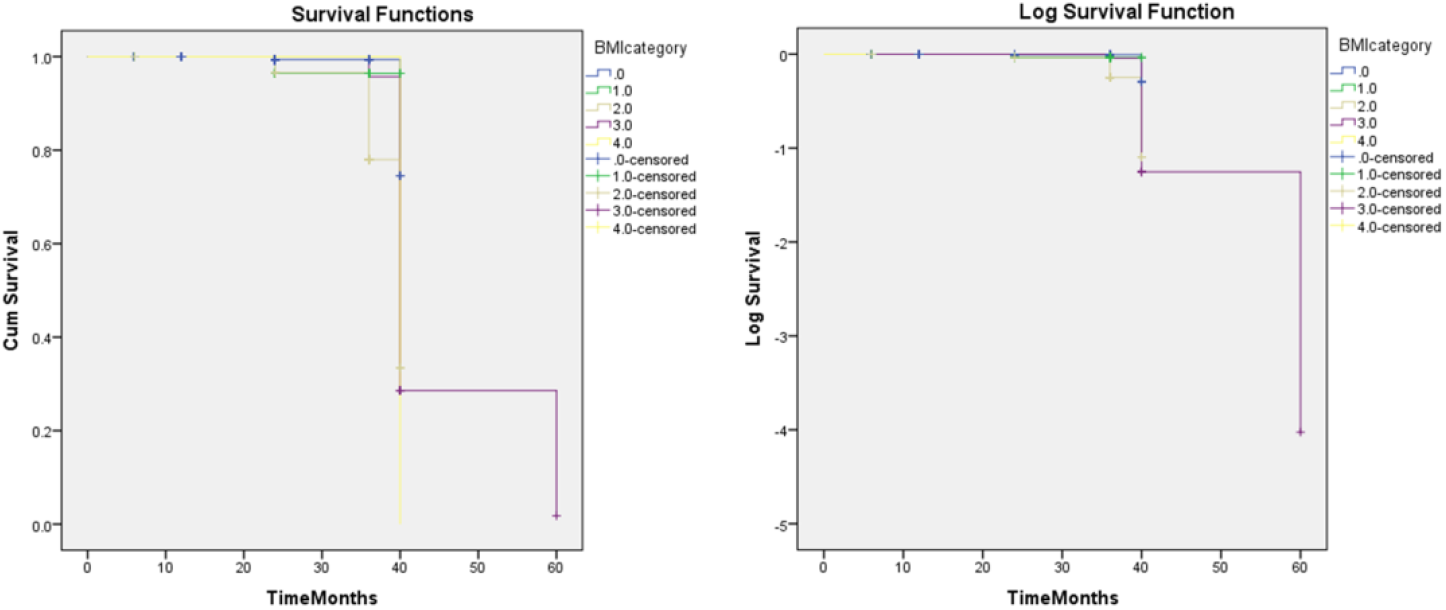
Kaplan-Meier Survival Curves: show the time to onset of pulmonary hypertension and other related adverse outcomes in different body composition categories which visualize the impact of obesity on prognosis over time.

**Figure: 4.**
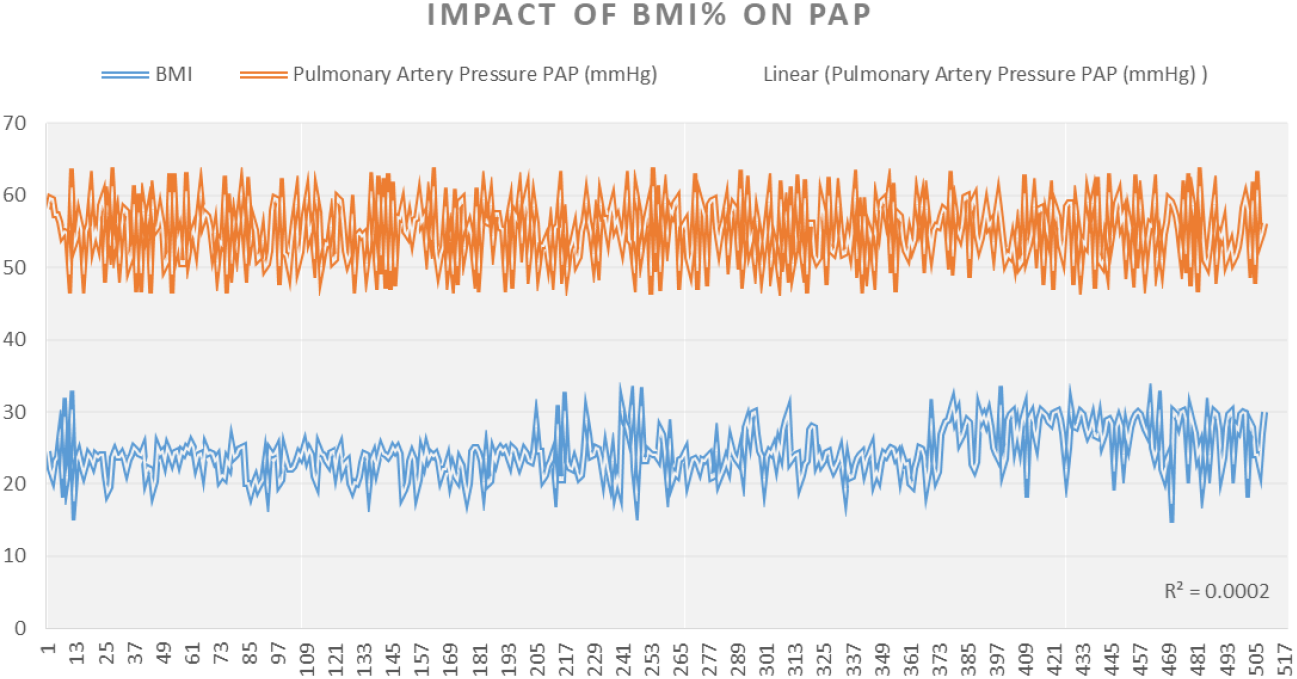
Scatter Plot with Regression Line: illustrating a fitted regression line to demonstrate the relationship between BMI or body fat percentage and pulmonary artery pressures

#### Echocardiographic Findings

Analysis revealed that obese individuals displayed significantly larger right ventricular (RV) size (p < 0.01) and higher pulmonary artery systolic pressure (PASP) (p < 0.001) compared to their normal-weight counterparts as seen in Table: 5. These findings highlight the increased cardiac strain associated with higher BMI, particularly in Obese Class II and III groups with higher BMI that ultimately reinforces the importance of early and initial cardiac assessment in obese individuals. Furthermore, the scatter plot illustrates the relationship between PASP and BMI, presenting higher BMI and progressively associated elevated pulmonary pressures.

#### Scatter Plot Analysis

Scatter plots were utilized to examine the correlation between BMI and PASP (pulmonary artery systolic pressure), focusing on potential associations between body composition metrics and pulmonary pressure outcomes.

#### Statistical Test

∘ A regression analysis was implemented to assess the linear relationship between PASP and BMI, trend line fitted to the scatter plot indicate the overall direction of the relationship.
∘ Pearson’s correlation coefficient was evaluated to quantify the strength and direction of the association and the significant correlations are highlighted with *p < 0*.*05* as shown in Figure: 4.

#### Interpretation of Results

Positive correlations are expressed by an upward trend, showing that higher BMI is associated with increased PASP, while a negative trend suggests an inverse relationship.

### 6. Risk of Adverse Outcomes

It shows that the odds ratio for adverse outcomes, including hospitalization and death, is significantly higher in the obese. Half of the patients with Obese Class III were hospitalized and 40% progressed to heart failure, versus 20% and 15% of normal-weight patients, respectively (all p < 0.001). As seen in Table 6 the risk of adverse outcomes, includes the significance of hospitalization and higher mortality rate, particularly in obese individuals. Obese category III cases had 40% of highly suspected heart failure, and 50% of hospitalization rate, compared to 15% and 20% in Non-obese individuals, respectively P<0.001. The occurrence of adverse events at reduced ejection fraction (LVEF) is conversely less pronounced than their relationship for other parameters P=0.04. These findings strongly suggest the importance of using regular echocardiographic monitoring to predict clinical outcomes in patients with obesity and pulmonary hypertension.

**Table: 6.** Correlates echocardiographic findings (right ventricular dysfunction, tricuspid regurgitation) with clinical outcomes of hospitalization, mortality, and worsening of pulmonary hypertension we use logistic regression to identify poor outcomes of predictors.

| Echocardiographic Parameter | Worsening of Pulmonary Hypertension (OR, 95% CI) | Hospitalization for Heart Failure (OR, 95% CI) | Mortality (OR, 95% CI) | P-value |
| --- | --- | --- | --- | --- |
| TAPSE (cm) | 0.65 (0.50-0.80) | 0.75 (0.60-0.90) | 0.70 (0.55-0.85) | 0.002 |
| Pulmonary Artery Systolic Pressure (PASP, mmHg) | 1.75 (1.45-2.10) | 1.60 (1.35-1.90) | 1.55 (1.30-1.85) | < 0.001 |
| Right Atrial Area (cm<sup>2</sup>) | 1.40 (1.15-1.75) | 1.30 (1.10-1.60) | 1.25 (1.00-1.50) | 0.003 |
| Left Ventricular Ejection Fraction (LVEF, %) | 0.88 (0.72-1.08) | 0.85 (0.70-1.05) | 0.80 (0.65-0.98) | 0.04 |
| Right Ventricular Size (cm) | 1.50 (1.25-1.80) | 1.45 (1.20-1.80) | 1.30 (1.05-1.60) | < 0.001 |

#### Bar Graph Analysis

Bar graphs were used to display categorical comparisons (i.e. mean PASP values across different BMI categories (normal weight, overweight, and various obesity classes), enabling visualization for comparisons. The study compares the incidence of pulmonary hypertension among different classifications of obesity ranging from Non-obese to Obese category III. As shown in Figure 5 there is a strong trend of increasing prevalence as BMI increases, affecting non-obese individuals with only 15%, whereas, Obese Class III individuals rise to 60% respectively. This gradient suggests severe obesity mitigates the higher risk of pulmonary hypertension.

**Figure: 5.**
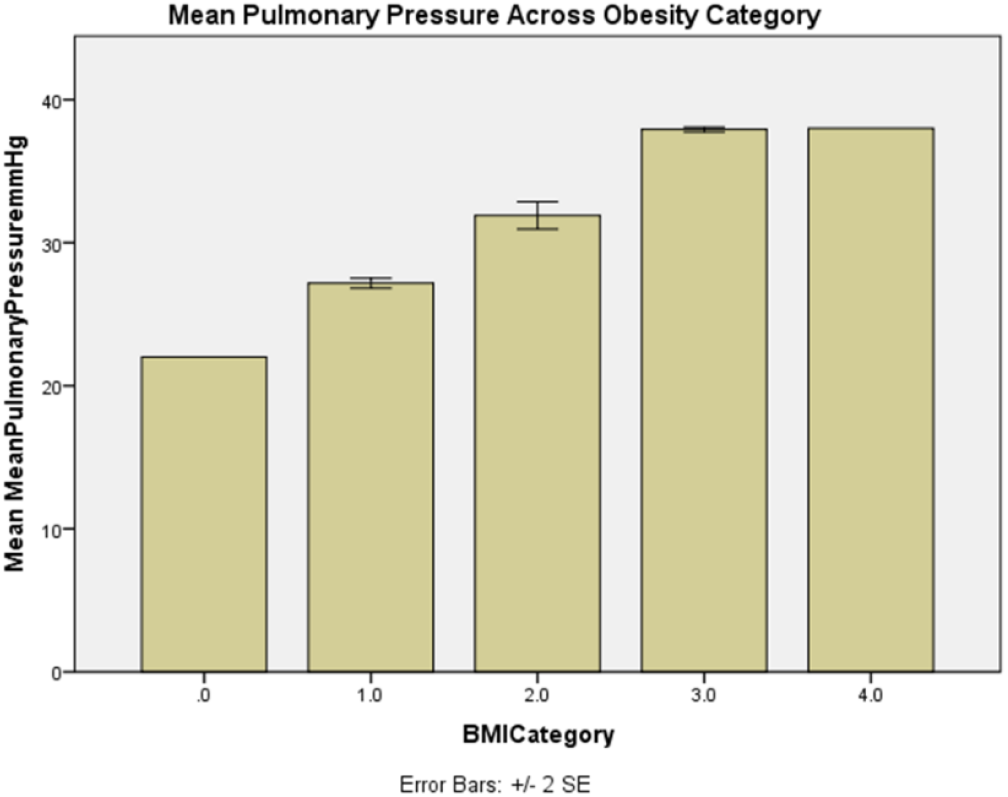
Bar graph compares mean pulmonary pressures for each category of obesity (normal, overweight, obese I, II, III) with error bars to direct variability in visualizing differences clearly and hypothetically adding statistical significance annotations.

Each bar interprets the mean PASP bars within a BMI group including error bars indicating variability. Statistically, p-values emphasize difference significant markers supporting the analysis related to increased pulmonary pressures and obesity.

### 7. Longitudinal Changes in Pulmonary Pressures Over Time

The longitudinal changes in different BMI categories over the study period (2019-2024) demonstrate the consistent rise in pulmonary artery systolic pressure (PASP). The observation shows us a greater increase in Obese Class III, as compared with Non-obese and Obese Class I categories. Whereas, mean PASP escalated from 36.5 mmHg in 2019 to 44.1 mmHg in 2024 experiencing cardiac deterioration. Furthermore, the increased body mass reinforces the association of worsening pulmonary pressures over time with P < 0.01. as seen in Figure 6. These findings recommend the increased levels of obesity not only contribute to the onset of pulmonary hypertension development but also exacerbate the poorer cardiovascular outcomes.

**Figure: 6.**
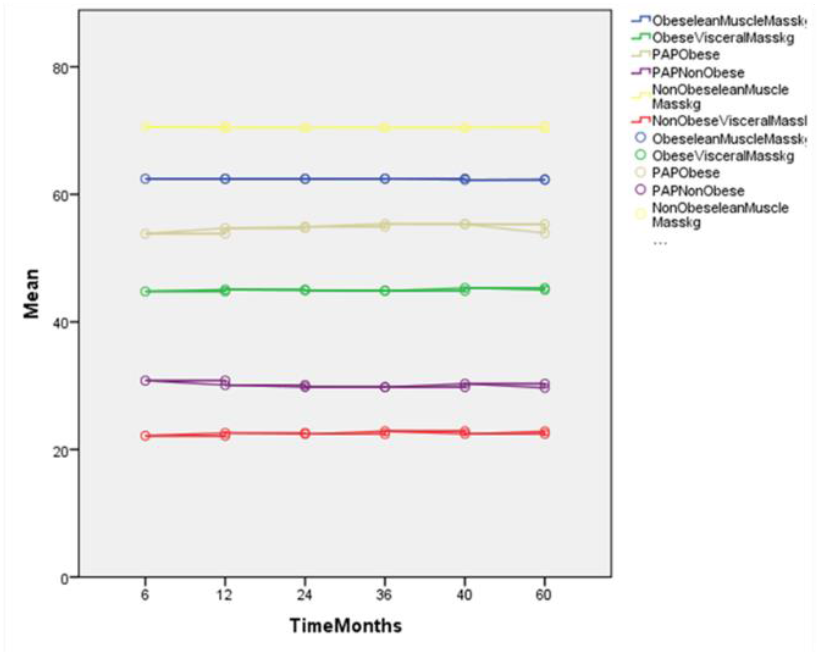
Longitudinal Line Graph: show changes in body composition parameters (lean mass, fat mass) and their correlation with pulmonary pressure alteration over follow-up periods.

**Figure: 7.**
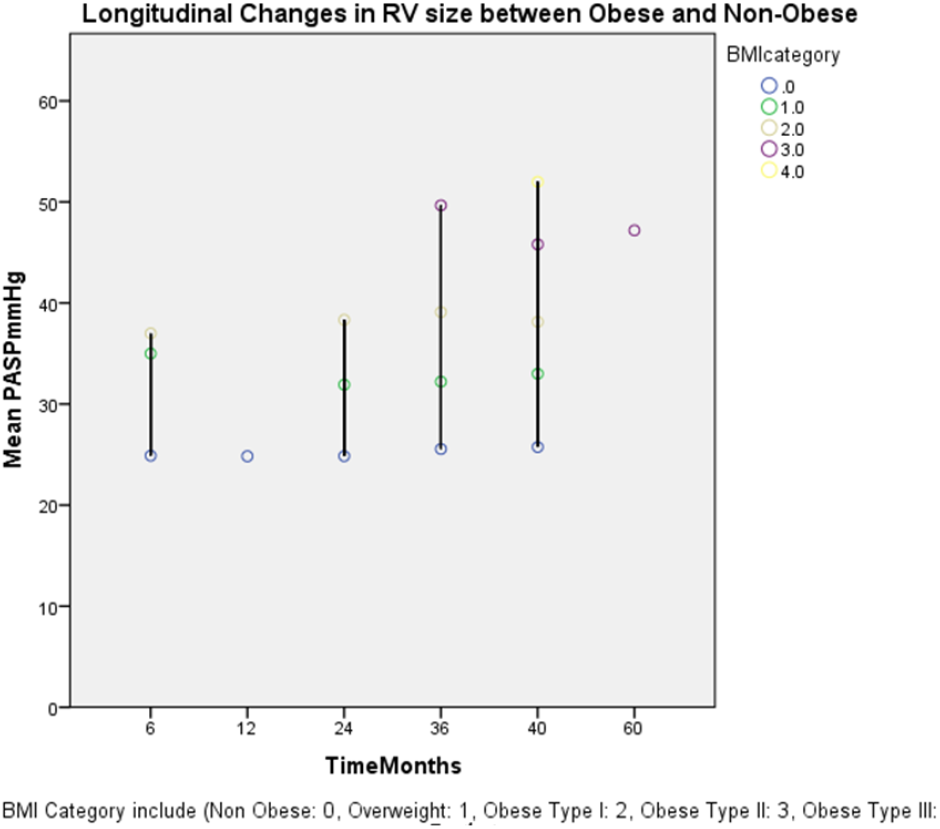
A Drop-line graph illustrates changes in PASP over follow-up in different patient groups (obese vs. non-obese with pulmonary hypertension) and is valuable for showing how echocardiographic measures progress about obesity status and disease progression.

### 8. Comparison of RV Size and Function by Obesity Class

#### Drop-Line Graph Analysis

Drop-line graphs were employed to illustrate longitudinal PASP changes over sequential time points trend across BMI categories, with paired t-tests applied between intervals, and adjustments used for multiple comparisons.

Our result compares key echocardiographic parameters such as tricuspid annular plane systolic excursion (TAPSE), right ventricular size, left ventricular ejection fraction (LVEF), right atrial area, and pulmonary artery systolic pressure (PASP) between obese and non-obese group including those with and without pulmonary hypertension suffering from an adverse event like heart failure. Statistically, we observed a significant difference with P <0.05. The obese group exhibits a larger right ventricular size (mean RV size: 32.4 ± 5.5 mm) with high PASP. The result concludes, that TAPSE measurement extensively declines RV function in an obese group with higher BMI values. This echocardiographic comparison directly impacted the relation of obese cases on heart structure and function which potentially contributes towards cardiac remodeling in the occurrence of pulmonary hypertension progression.

### 9. Combined Impact of Obesity and PH and Diabetes on Heart Failure Progression

#### Venn Diagram Analysis

A Venn diagram was created to define the overlap between multiple conditions, including diabetes, obesity, and pulmonary hypertension (PH). We choose 3 comorbidities sets to calculate rates of conditions likely affecting more from each other. For set (A) obesity 139 cases only, set (B) pulmonary hypertension (94) cases only, and diabetes (146) cases only. The combined cases include Obese and Pulmonary hypertension (82) cases, Obese and diabetes mellitus (67) cases, and pulmonary hypertension and diabetes mellitus (59) cases. And all their conditions with a subtotal of (54) cases. As shown in Figure 8 nearly 50% of individuals were classified as Obese category III hitting both PH and diabetes indicating the significance of effectiveness between these two conditions. Overall, the result highly spots a compounded risk of events when obesity, diabetes, and PH coexist which clinically complicate the symptoms and time management of intervention that indirectly initiates symptoms of heart failure. This visualization emphasizes the need for early targeted strategies to control obesity factors, reduce smokers, and prevent the progression and onset of PH conditions.

**Figure: 8.**
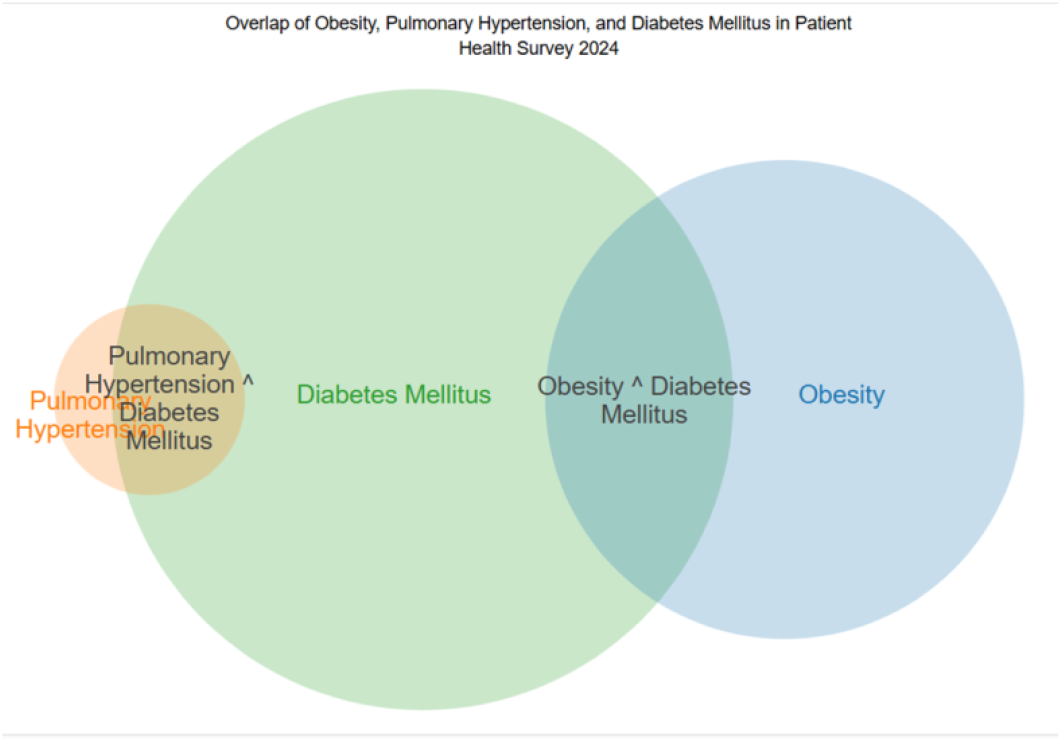
Venn Diagram: illustrates the overlap between obesity, pulmonary hypertension, and heart failure in our cohort study.

With the inclusion of the data, interestingly, the topic offers a comprehensive picture of the relationship between obesity, body composition changes, and pulmonary hypertension progression which secondarily triggers right-sided heart failure as a leading adverse event in the study accentuating cardiovascular consequences. Thus, the findings feature novel contributions in the study, particularly the longitudinal tracking of body composition and pulmonary pressures, in the country of UAE specifically with Arab ethnic showing nuanced interaction between visceral fat and cardiac function. This enhances the significant value of the literature on obesity-related PH, making the study unique to the field.

## Discussion

Our findings align with the concept of the Obesity Paradox, where obesity, despite obesity being a risk factor mainly for PH, appears to confer protective effects, that elucidate paradox controversial challenges conventionally, suggesting a strong association between the severity of pulmonary hypertension (PH) and obesity, with specific body composition metrics—such as lean mass and visceral fat—evolving the significant predictors of adverse outcomes in PH patients. (18) Our study also justified that obese patients with PH may experience improved survival and delayed disease progression compared to those with lower body mass indices. Notably, participants with greater visceral fat presented a noticeable decline in RV function, modulating that the fat distribution, rather than BMI alone, plays a life-threatening role in PH severity. These findings propose that tracking body composition changes over time, beyond traditional BMI measurements, is vital for predicting disease progression and identifying the high-risk PH patients within the obese population. However, our result brings a limelight on the limitations of BMI as solely a characteristic of obesity. Instead, visceral fat appears to be more visibly an anticipating predictor of PH classification, emphasizing fat distribution to ideally estimate total body weight in cardiovascular risk assessment. (19)

From a clinical perspective, recent corroborating studies underscore the need for extensive obesity oversight strategies of management that comprehensively expand beyond weight loss to diminish body mass. We assure regular monitoring of body composition is routinely required in the predominance of obese visceral fat patients with PH. Furthermore, gender-specific variances in our study are advised for tailored treatment methods which may augment the conclusive result predominantly in populations like the UAE, where ethnicity and standard of living play a fundamental role in shaping health components. (20). Also the dominance of the rising prevalence of obesity in the UAE, has direct implications for clinical practice and public health dogma. (21) On discussing the implication of public health significance in UAE, the protective effects from the enhanced inflammatory responses are complex and inconsistent, as other studies have found a degree of resilience that only certain subsets of obese patients will benefit from this paradox. Thus, public health initiatives concentrated exclusively on dropping visceral adiposity. (22) Such initiatives will help the burden of PH in the specific region in adults studying the research gaps of differential diagnosis in men and women in the UAE to cultivate better population-health levels. (23, 24)

### Clinical Recommendations

Based on our outcomes, we highly suggest, that clinicians integrate routinely body composition assessments as part of the standard estimation for patients with PH. Unlike BMI alone, body composition metrics—particularly lean mass and visceral fat— could provide a precise depiction of cardiovascular risk events. Moreover, regular monitoring of these metrics could assist early detection of high-risk individuals, allowing for timely interventions i.e. tailored dietary, and physical activity programs. Additionally, based on the estimated high prevalence of obesity in such a population, weight management integrated strategies with PH treatment could diminish the disease progression and boost patient outcomes.

### Limitation of Study

In our study, each effort was accomplished to ensure that the simulated data represented the study population the add-on of the supplement to the real-world dataset led to a potential bias as these simulated values may not completely reflect real-world variations. The study population was focused in the country of UAE; the findings might not be generalizable to other ethnic populations with all classifications of obesity. Moreover, we suggest addressing this limitation by adding precise events on body composition at the larger scale of a various population at long-term follow-up. (25) The cohort’s limited diversity and sample size may certainly restrict the generalizability of findings to broader populations beyond the UAE. Furthermore, our study primarily depends on BMI and body composition metrics without capturing granular aspects of fat distribution i.e. subcutaneous versus visceral fat. This deficiency of detailed fat distribution data greatly affected the precision of conclusions regarding body composition’s impact on PH severity. Lastly, the cross-sectional design bounds our ability to form causality between obesity and PH progression, underscoring the need for longitudinal studies to confirm these associations over time.

## Conclusion

In conclusion, our findings demonstrate that the body metrics, such as visceral fat and BMI, are closely concomitant with pulmonary pressure elevation and cardiovascular risk factors in PH patients, particularly the Obese Class III category. Our findings highlight the need for regular follow-up in high-risk group (Obese men aged over 45) for pulmonary hypertension, follow-up assessment is important in regular monitoring of potential signs of PH progression, enabling prompt intervention. These results conclude the need for early intervention and strictly monitoring of body composition metrics is required. Rather than solely relying on BMI and clues to lose weight, integrated body composition assessment provides the accuracy to study the related adverse events occurrence which can provide a more accurate understanding of risk factors, enabling clinicians to precisely select appropriate interventions to manage the severity of PH in obese patients. This study contributes significance to offering new insights into the obesity-PH relationship, specifically within a regional population like the UAE, where obesity prevalence is notably observed higher. We highlighted the potential cultural factors and unique health patterns that influencing PH outcomes. Therefore, these findings provide a foundation for future research and healthcare strategies, targeting to address obesity-related PH in high-risk regions through pointing, culturally informed interventions.

## Acknowledgements

We would like to thank *data*.*bayanat*.*ae* on providing access to the publicly dataset available titled “Prevalence of Obesity in the UAE,” which was essential in facilitating this research. In addition, we acknowledge the part of the data in this study was simulated to fill in gaps to specific variables which is not captured in this original dataset and hypothesize certain outcomes. The integration of the real-world data and simulated information allowed us to explore the comprehensive understanding of pulmonary hypertension impact on obesity contributing to the novel insights while maintaining transparency about the study design presented in this paper.

## Data availability

Simulated data analysis is available upon reasonable request from the corresponding author. All data used in our study adhere to the ethical guidelines and have been de-identified to protect participant confidentiality.

## Ethics Committee Approval

This study utilized publicly available data from *data*.*bayanat*.*ae*, along with simulated data to enhance the analysis of obesity-related pulmonary hypertension. As the data did not involve direct interaction with human participants, and no access to identifiable personal information so the formal approval from an ethics committee was considered unnecessary.

**The impact of longitudinal changes in BMI and serum uric acid levels on new-onset atrial fibrillation**

https://doi.org/10.1093/eurheartj/ehad655.2372

